# Neuroimaging access in Chad: Infrastructure barriers and opportunities for accessible magnetic resonance imaging

**DOI:** 10.64898/2026.09.10.26362077

**Authors:** Israa Hissein, Jingting Yao, Foksouna Sakadi, Esias Bedingar, Ming Zhao, André J.W. van der Kouwe, Jerome L. Ackerman

**Author notes:** These authors made equal contributions (Israa Hissein, Jingting Yao). These authors are co-senior authors (André J.W. van der Kouwe, Jerome L. Ackerman). Corresponding author: André J.W. van der Kouwe.

## Abstract

Limited access to neuroimaging remains a major constraint on neurological care in many low-resource areas. We conducted a descriptive landscape analysis of neuroimaging access in Chad using hospital-reported data, publicly available healthcare and infrastructure data, and contextual information from clinicians and radiology personnel at major hospitals in N’Djamena. This analysis characterizes key constraints on neuroimaging access in Chad and highlights practical considerations for expanding imaging capacity in resource-limited settings. Chad has only one magnetic resonance imaging scanner for a population of approximately twenty-one million. The country’s major hospitals and imaging workforce are concentrated in the capital, and the sole MRI system is an older 0.35 T scanner located at a private hospital. Access is further limited by scan cost, geographic centralization, unreliable electricity, limited technical support, and shortages of trained imaging personnel. Hospital data indicate a substantial burden of neurological conditions, particularly stroke and neurological infections, for which imaging can be important for diagnosis and management. We place these findings in the context of emerging lower-cost, compact, and mobile magnetic resonance imaging technologies. Such systems may reduce some infrastructure and siting requirements, but limitations related to image quality, maintenance, workforce capacity, and long-term sustainability remain important considerations.

## 1. Introduction

Neurological disorders constitute a substantial health burden across sub-Saharan Africa (SSA), including stroke, cerebral malaria and other central nervous system infections, epilepsy, and traumatic brain injury (Samba, 2001; Logroscino et al., 2024; Sakadi, Dakissia, et al., 2021). Cerebral malaria alone affects more than 600,000 people in SSA annually, predominantly young children, and survivors may experience persistent neurological complications (Idro et al., 2010; Trivedi and Chakravarty, 2022; Abdelmalik and Kashbour, 2024; Sakadi et al., 2024). Accurate diagnosis and management of many neurological conditions depend on medical imaging, yet access to advanced imaging remains highly unequal. Approximately 65,000 magnetic resonance imaging (MRI) systems are installed worldwide— about 7 scanners per million people—with much of this capacity concentrated in high-income countries and limited access across many low-resource settings (Organisation for Economic and Development, 2022).

The disparity is particularly pronounced in Chad, a landlocked country in north-central Africa with a population of approximately 21 million (World Bank, 2023; Chad country brief, 2023). Despite its large geographic area and substantial neurological care needs, Chad currently has only one MRI scanner (Sakadi and Mateen, 2018; Agbetou et al., 2023). Major hospitals, neurologists, radiologists, and imaging services are concentrated in the capital, N’Djamena. Limited imaging capacity occurs alongside broader infrastructure constraints: less than12% of the population has access to electricity, and unreliable power, transportation barriers, equipment maintenance, and shortages of specialized personnel further complicate the operation and expansion of conventional MRI services (Kindzeka, 2022; Moyo et al., 2023; Bamisile et al., 2023).

Conventional MRI systems impose substantial financial and infrastructural requirements, including high acquisition and maintenance costs, stable electrical power, cooling systems, specialized installation space, and trained technical personnel (Liu et al., 2021). Recent developments in compact, low-field, portable, and lower-cost MRI systems have therefore generated interest as potential approaches for expanding neuroimaging access in resource-limited settings (Geethanath and Vaughan, 2019; Parasuram et al., 2023; Yuen et al., 2022; Beekman et al., 2022; Cooley et al., 2021; Huang et al., 2018; Sheth et al., 2020; Arnold et al., 2023; Obungoloch et al., 2023). Some of these systems reduce requirements for power, cooling, shielding, or dedicated installation space (Cooley et al., 2021; Liu et al., 2021; Lau et al., 2023). However, reduced infrastructure requirements do not necessarily translate into practical or sustainable clinical implementation. Their usefulness depends on local clinical needs, diagnostic performance, affordability, maintenance capacity, and availability of trained clinical and technical personnel.

Here, we characterize neuroimaging access in Chad, with a focus on neurological care and imaging resources, major constraints on MRI access, and the potential relevance of emerging accessible MRI technologies.

## 2. Methods

We compiled hospital-reported statistics, publicly available healthcare and infrastructure data, and contextual information obtained through open professional communications with neurologists, radiologists, and hospital personnel in Chad. Hospital data included inpatient neurological cases and imaging-resource information from major hospitals in N’Djamena. Publicly available sources were used to compile demographic, healthcare, infrastructure, and MRI availability data for Chad and other African countries. Published literature was additionally used to place the identified barriers in the context of emerging low-cost, compact, and low-field MRI technologies.

Hospitalization data were reported as provided by participating institutions; counts covering periods shorter than 12 months were annualized for comparison. Contextual communications were informal and were not conducted as structured interviews or surveys. No identifiable patient information was accessed or used.

## 3. Results

### 3.1 Neurological care and hospital-reported disease burden

Hospital-reported data from the National Reference Teaching Hospital demonstrate a substantial burden of neurological disease, with stroke accounting for the majority of neurological hospitalizations between 2023 and 2025 (**Fig. 1**). Stroke represented 192 of 254 inpatient neurological cases (75.6%) in 2023, 259 of 326 cases (79.4%) in 2024, and 300 of 356 cases (84.3%) in 2025. Ischemic stroke was the most frequently reported stroke subtype in each year, followed by hemorrhagic stroke (**Fig. 1D**). Other reported neurological conditions included cerebral infection, epilepsy, intracranial expansion, medullary compression, encephalopathy, peripheral neuropathy, and lumbosciatica.

**Fig. 1.**
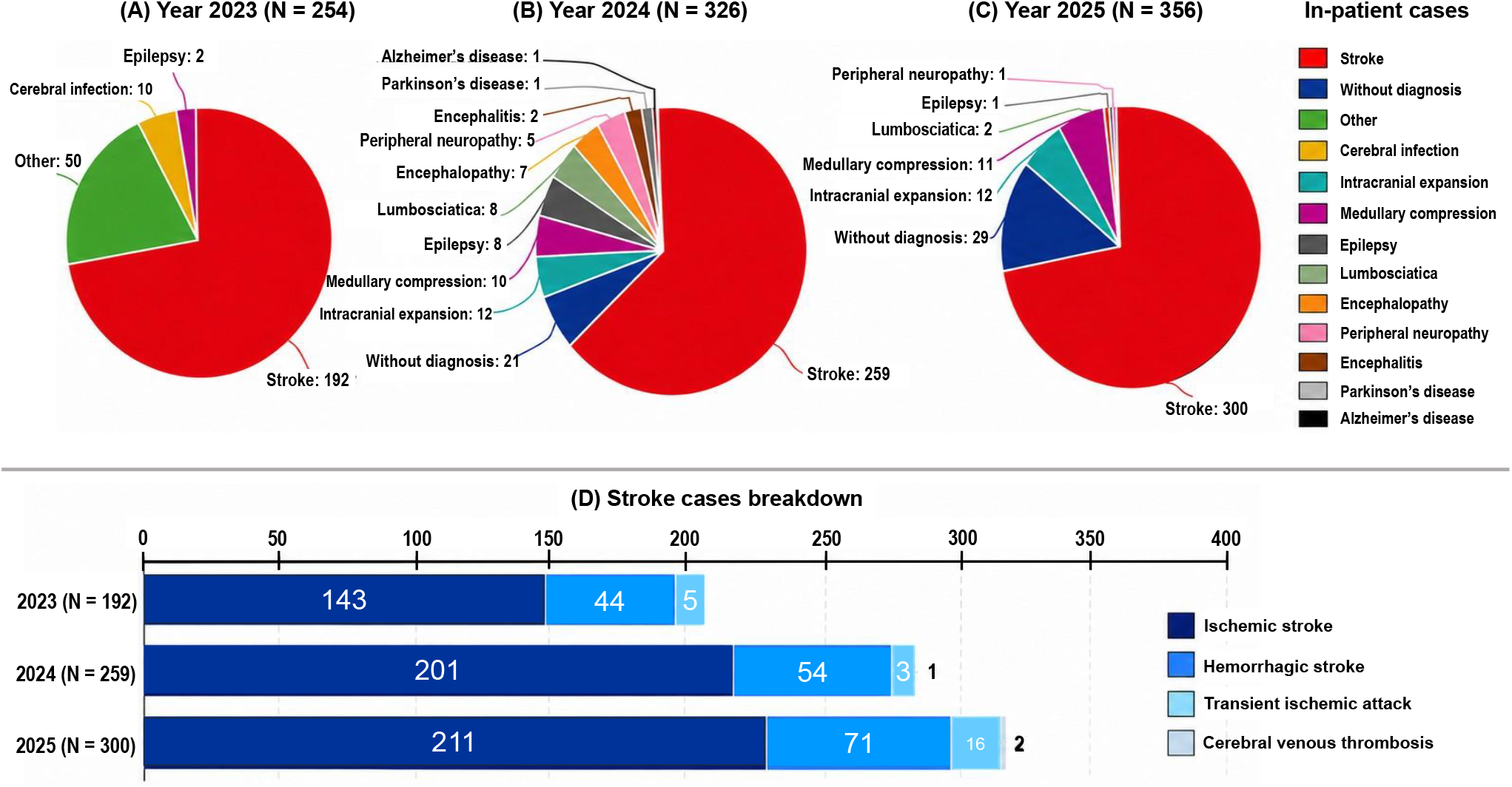
Distribution of inpatient neurological cases and stroke subtypes, 2023–2025. **(A–C)** Hospital-reported inpatient neurological cases in 2023, 2024, and 2025, respectively, categorized by diagnosis. **(D)** Distribution of reported stroke cases by subtype, including ischemic stroke, hemorrhagic stroke, transient ischemic attack, and cerebral venous thrombosis.

At the National Reference Teaching Hospital, hospitalization refers to patients who remained in the hospital for at least three days and had a documented hospitalization record. Clinicians indicated that limited resources and delays in diagnostic testing or treatment can prolong hospital stays, while hospitalization is generally reserved for more complicated cases. Consequently, more than 90% of neurological patients are managed without hospitalization. Although the exact number of outpatient visits to the Neurology Department was not available, clinicians reported more than 50 patient consultations per day, indicating that inpatient data capture only a portion of the overall neurological caseload.

Although epilepsy accounted for only a small proportion of the inpatient cases shown in **Fig. 1**, most epilepsy cases are managed on an outpatient basis, and clinicians report a substantial volume of such patients. Previous studies have similarly documented the burden of epilepsy and barriers to epilepsy care in Chad (Sakadi et al., 2024; Sakadi, Madjirabe, et al., 2021). Epilepsy may also be under-recognized when it occurs in association with another primary diagnosis, such as infection or stroke (Sakadi et al., 2024). During free neurological consultations in rural areas, Chadian neurologists have estimated a high prevalence of epilepsy in some southern communities, particularly among younger individuals. Financial barriers remain an important limitation to hospital-based diagnosis and treatment of neurological conditions (Sakadi et al., 2023).

Additional hospitalization data were available from Renaissance Teaching Hospital and Sino-Chadian Friendship Hospital (**Table 1**). At Renaissance Teaching Hospital, neurological diseases accounted for 9% of all hospitalizations in 2023. The Sino-Chadian Friendship Hospital primarily provides general medical care and does not have a dedicated neurology department; patients with suspected neurological conditions are therefore often referred to hospitals providing specialized neurological care. Hospital-level data were incomplete because of differences in record-keeping and data-sharing practices, and some institutions were unable or hesitant to provide detailed statistics. The reported counts should therefore be interpreted as selected available hospital data rather than as a comprehensive estimate of neurological disease burden across Chad.

**Table 1.** Hospitalizations for selected neurological conditions at two major hospitals in Chad.

| Hospital | Period | Neurological condition | No. of cases |
| --- | --- | --- | --- |
| Renaissance Teaching Hospital | Jan 2022–Jun 2023 | Hemorrhagic stroke | 218 |
|  |  | Ischemic stroke | 452 |
|  | Jan 2024–Dec 2024 | Stroke | 167 |
|  |  | Epilepsy | 181 |
| Sino-Chadian Friendship Hospital | Jan 2023–Dec 2023 | Cerebral hematoma | 53 |
|  |  | Ischemic stroke | 47 |

Note: Case counts represent hospital-reported data for the indicated periods, which varied according to data availability. At Renaissance Teaching Hospital, stroke was reported as a combined category in 2024, whereas subtype-specific stroke data were available for other reporting periods.

### 3.2 Neuroimaging capacity and workforce in Chad

Chad’s major hospital-based neurological and radiological services are concentrated in N’Djamena. The four major hospitals considered in this study are the National Reference Teaching Hospital (CHU de Référence Nationale), the country’s largest public hospital; Renaissance Teaching Hospital (Hôpital de la Renaissance), a private hospital established in 2013; the Mother and Son Teaching Hospital (Hôpital de la Mère et de l’Enfant), which provides care primarily for women and children; and the Sino-Chadian Friendship Hospital (Hôpital de l’Amitié Tchad-Chine). At the time of data collection, Renaissance Teaching Hospital housed the only MRI scanner identified in Chad, consistent with previous reports of extremely limited MRI availability in the country (Sakadi and Mateen, 2018).

The scanner at Renaissance Teaching Hospital is a refurbished Siemens MAGNETOM C! 0.35 T system dating to 2005 or earlier. Clinicians reported limitations in image quality that can complicate interpretation, particularly when findings are subtle. MRI is also substantially more expensive than other commonly available imaging services. An MRI examination without contrast costs approximately US$224, compared with approximately US$100 for computed tomography (CT) scans. Costs for other hospital services are higher at the private Renaissance Teaching Hospital: admission costs approximately US$30 compared with US$10 at the public National Reference Teaching Hospital, while a specialized consultation costs approximately US$18 compared with US$6 at the National Reference Teaching Hospital. These cost differences further limit access to MRI for patients with limited financial resources. Patients with greater financial means may instead travel to countries such as Sudan, Cameroon, Egypt, Morocco, or Tunisia for MRI examinations.

The National Reference Teaching Hospital, despite being the country’s largest public hospital and a major center for neurological care, does not have an MRI scanner. It has two CT scanners, although only one was operational as of March 2025. The Radiology Department performs approximately 20–40 CT examinations per weekday, with weekends generally reserved for emergency cases (**Fig. 2**). In the absence of MRI, neurological evaluation relies primarily on clinical examination, laboratory testing, electroencephalography, and CT imaging.

**Fig. 2.**
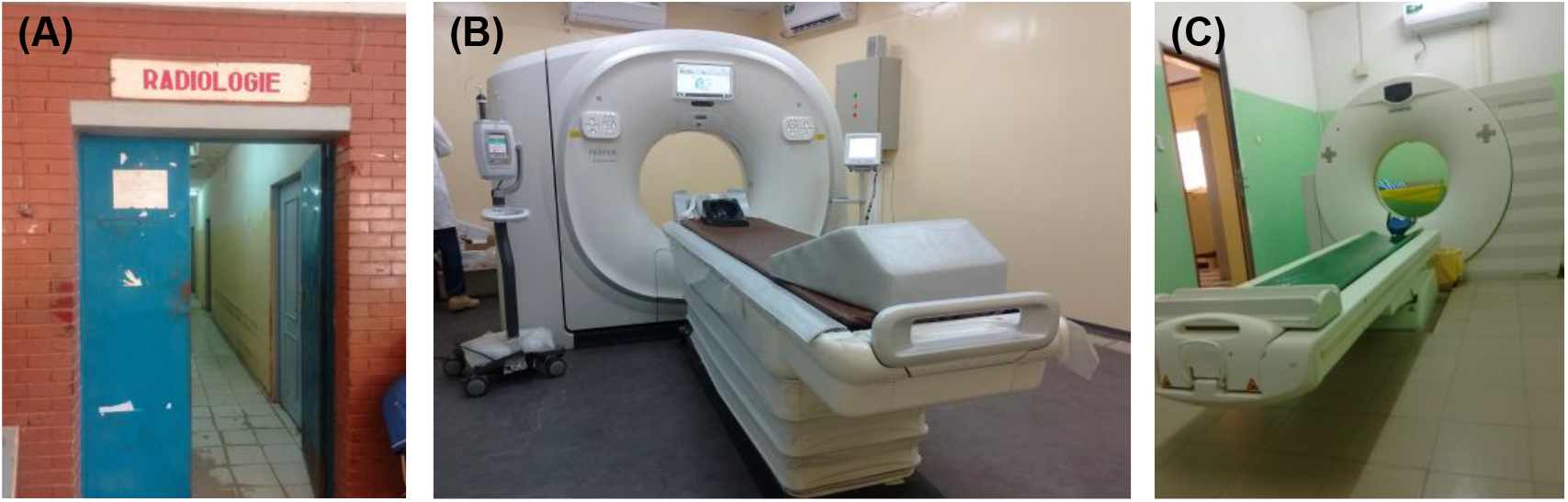
Radiology Department at National Reference Teaching Hospital, N’Djamena, Chad. The hospital lacks an MRI scanner but operates two CT scanners, which perform 20 to 40 scans per day on weekdays. Weekends are reserved for emergency cases. **(A)** Entrance to the Radiology Department, a gateway to essential diagnostic services. **(B)** A Siemens SOMATOM Emotion CT scanner, one of the two machines managing the weekday scan workload. **(C)** A FujiFilm CT Scenaria View scanner, the second operational unit in this department. Photographed in November 2023.

Specialized imaging and neurological personnel are also limited and geographically concentrated. As of April 0, Chad had eight neurologists, all based at the four major hospitals in N’Djamena; two were women. The neurologists meet weekly to review complex cases collaboratively. Ten radiological technologists were identified across the major hospitals: four at the National Reference Teaching Hospital, three at Renaissance Teaching Hospital, and three at the Sino-Chadian Friendship Hospital. As of December 2025, the National Reference Teaching Hospital alone was staffed by three neurologists, four radiologists, and four radiological technologists. The concentration of both imaging equipment and specialized personnel in the capital further limits access for patients living outside N’Djamena.

### 3.3. MRI availability in Chad within the African context

MRI availability remains limited across much of Africa, although substantial variation exists between countries (**Table 2**). We compiled available estimates of MRI units from the World Health Organization and published sources, with corresponding population estimates obtained from the World Bank (World Bank, 2023). Because the most recent available data vary by country and some sources report only public- or private-sector scanners, these estimates should be interpreted as an approximate comparison of MRI availability rather than a precise contemporaneous census.

**Table 2.**
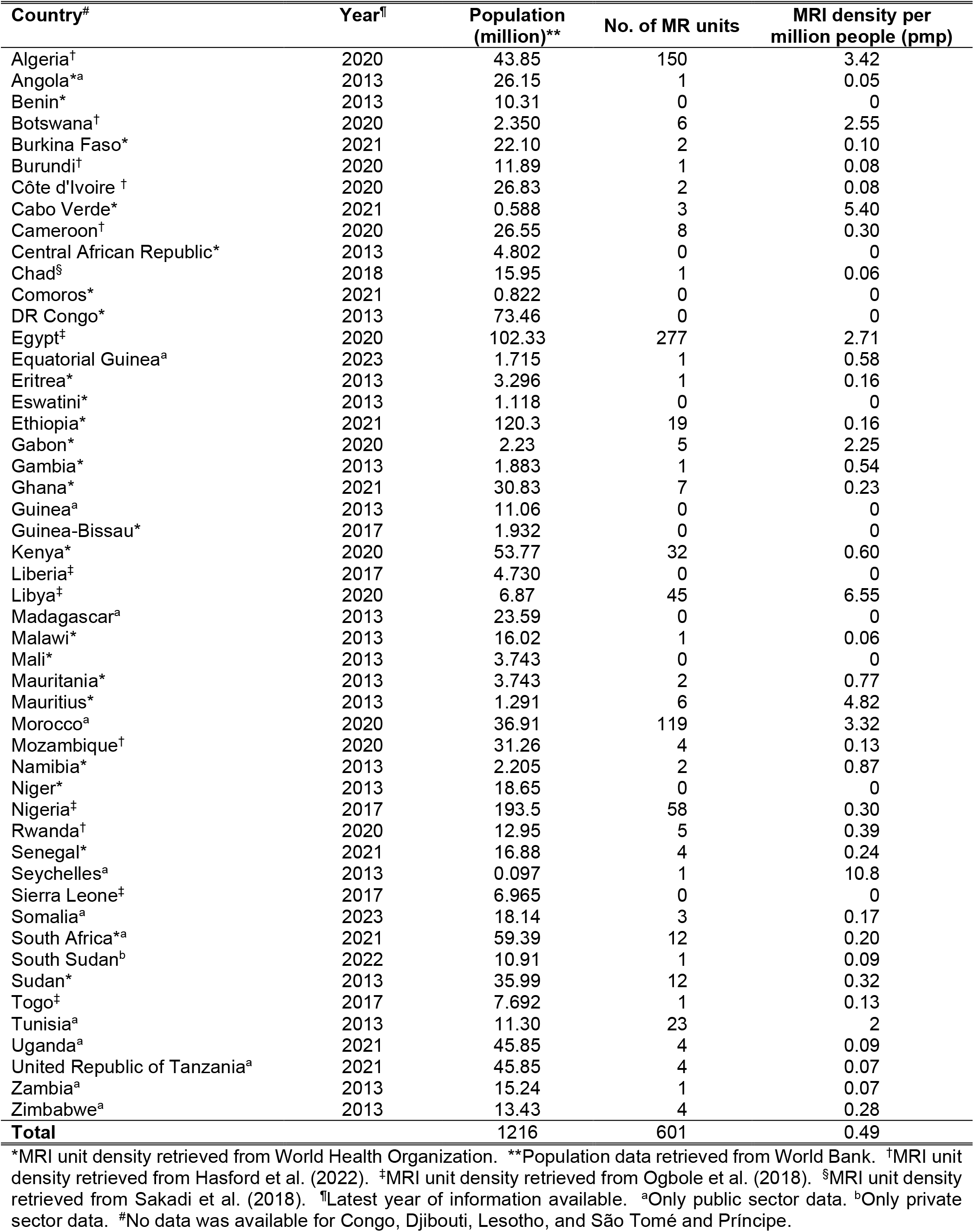
Distribution of MRI units in African countries.

Within the compiled dataset, Chad had one reported MRI unit in 2018, corresponding to approximately 0.06 units per million people, consistent with previous reporting of highly limited MRI availability in the country (Sakadi and Mateen, 2018). This places Chad among the countries with the lowest reported MRI density in Africa (**Table 2**). Several countries had no reported MRI units, while others had substantially greater availability, including Algeria, Botswana, Egypt, Libya, Mauritius, Morocco, and Seychelles, each with more than two MRI units per million people in the available data. Nevertheless, MRI availability remained below one unit per million people in many of the countries represented in the dataset.

The contrast is even greater when Chad is compared with high-income areas. As illustrated in **Fig. 3**, Chad had approximately 0.06 MRI units per million people compared with approximately 37 per million in the United States (Organisation for Economic and Development, 2022). The disparity in scanner availability occurs alongside large differences in other healthcare and infrastructure indicators that may influence the ability to install, operate, and maintain MRI systems.

**Fig. 3.**
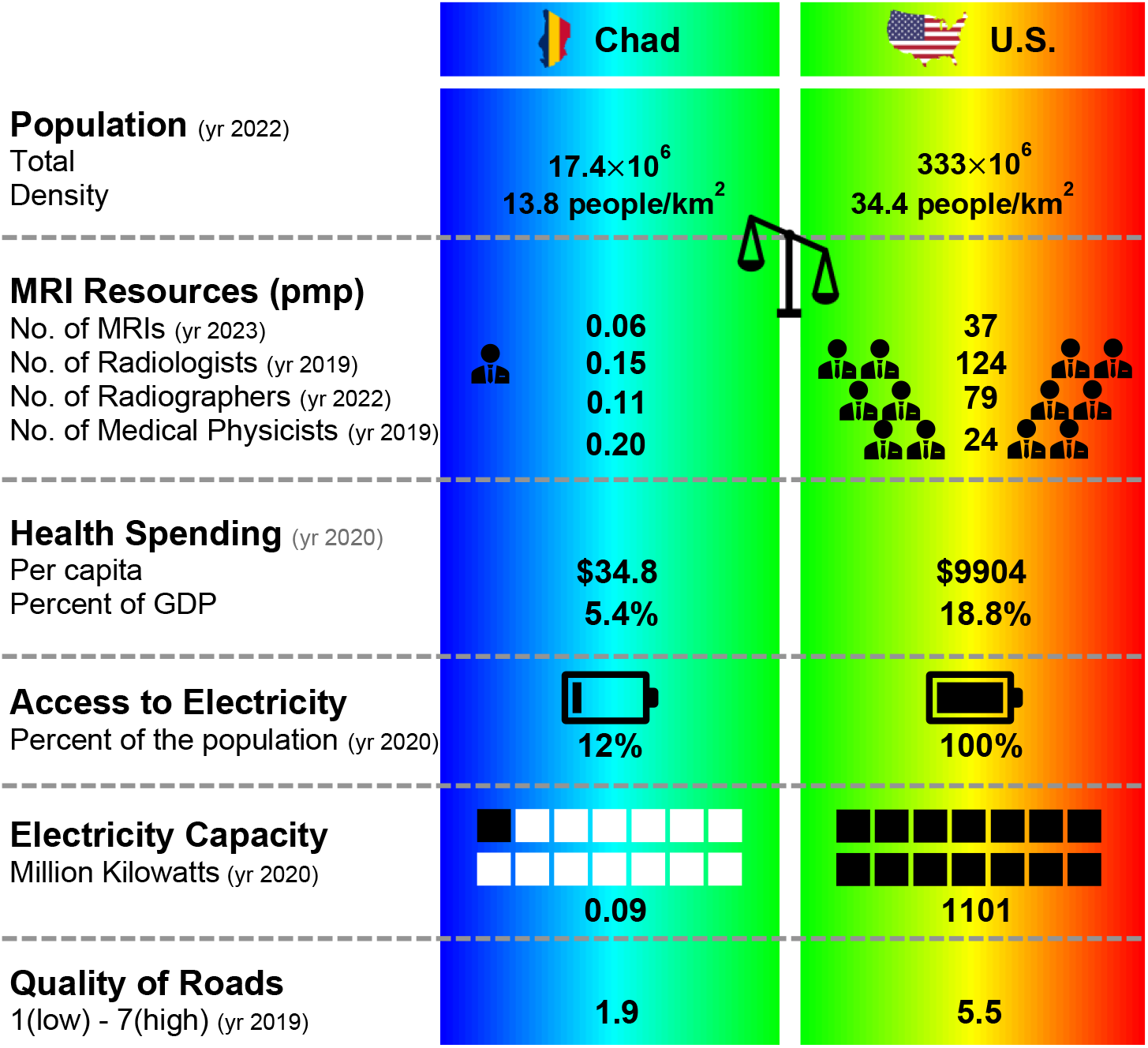
A Comparison of radiological resources in Chad and the U.S. Data Sources: World Bank, World Economic Forum, OECD, U.S. Energy Information Administration.

### 3.4. Infrastructure and operational barriers to MRI access

Limited transportation and electrical infrastructure represent additional barriers to neuroimaging access in Chad. Because major hospitals and specialized imaging services are concentrated in N’Djamena, patients living outside the capital may need to travel substantial distances for diagnostic imaging. Road conditions can make such travel difficult, with poorly maintained roads and seasonal flooding further limiting accessibility (Kindzeka, 2022; Moyo et al., 2023). During the rainy season, heavy rainfall can restrict road access, while limited road lighting and poor road conditions create additional transportation challenges (Kindzeka, 2022; Moyo et al., 2023; Sakadi and Mateen, 2018).

Reliable electricity is another important constraint. Electricity access remains limited in Chad, and the national electrical network is concentrated primarily in N’Djamena, where power outages and fluctuations remain common (Bamisile et al., 2023). These limitations are particularly relevant for conventional MRI systems, which require stable electrical power and substantial supporting infrastructure for routine operation (Liu et al., 2021).

Equipment maintenance and technical support present further challenges. MRI systems require routine quality assurance, preventive maintenance, and access to specialized technical expertise to maintain reliable performance (Epistatou et al., 2020). In Chad, limited local access to specialized service engineers and replacement parts can increase both the cost and time required for equipment maintenance and repair. In a setting with very limited imaging capacity, prolonged downtime of a single scanner can substantially reduce access to diagnostic imaging.

Detailed, publicly available information describing Chad-specific regulatory standards and quality-control procedures for MRI was limited. The Ministry of Public Health is responsible for oversight of medical equipment and healthcare regulation; however, specific information regarding MRI quality assurance requirements, enforcement, and implementation was not readily available. Together, limitations in infrastructure, technical support, workforce capacity, and quality-control resources present challenges not only to acquiring additional MRI systems, but also to maintaining reliable imaging services over the long term.

## 4. Discussion

### 4.1 Implications of the current landscape

The findings highlight a mismatch between neurological care needs and the capacity of the health system to provide timely and sustainable neuroimaging. Previous work has documented severe limitations in neurological resources in Chad (Sakadi and Mateen, 2018), and our data show that these limitations extend beyond scanner availability alone. Geographic concentration of services, affordability, workforce capacity, transportation, maintenance, and supporting infrastructure all influence whether imaging is practically accessible. Increasing the number of scanners without addressing these interdependent barriers may therefore have limited impact on access.

This distinction is particularly important when interpreting conventional measures such as MRI units per million people. Scanner density provides a useful basis for international comparison, but it does not capture whether imaging is geographically or financially reachable. In Chad, specialized neurological and imaging services are concentrated in N’Djamena, while poor road conditions and seasonal flooding can further impede travel from other regions (Kindzeka, 2022; Moyo et al., 2023). The country’s sole MRI capability is also located in a private hospital, whereas the major public referral hospital does not have MRI. Together with the relatively high cost of MRI, this suggests that availability and accessibility are not equivalent, a distinction that may also be relevant in other resource-limited settings. Financial barriers have similarly been identified as an important constraint on neurological and stroke care in Chad (Sakadi et al., 2023).

The neurological disease profile further argues against viewing expansion of imaging capacity as simply a question of acquiring more MRI systems. Stroke represents a major component of hospital-based neurological care, but CT remains essential for acute evaluation, particularly when rapid identification of intracranial hemorrhage is required. At the same time, the substantial burden of epilepsy, neurological infections, and other conditions indicates a need for imaging capabilities extending beyond acute stroke care (Sakadi et al., 2024; Sakadi, Madjirabe, et al., 2021). An appropriate imaging strategy should therefore be guided by locally important clinical questions and by how MRI can complement, rather than replace, existing CT services.

More broadly, these findings emphasize the difference between acquiring an MRI scanner and establishing a sustainable MRI service. MRI depends on reliable power, appropriate siting, trained personnel, quality assurance, maintenance, and access to technical support (Liu et al., 2021; Geethanath and Vaughan, 2019). In a setting where several of these supporting factors are constrained simultaneously, equipment acquisition alone may produce limited or short-lived gains. Expanding MRI access in Chad will therefore require attention to the entire imaging ecosystem, not simply the number or field strength of scanners.

### 4.2 Potential role of accessible MRI

In this context, we use “accessible MRI” to refer broadly to MRI technologies and approaches designed to reduce the financial, physical, infrastructural, and geographical barriers that limit patient access to MRI. The concept therefore extends beyond simply providing a lower-field version of conventional MRI. In Chad, this broader framing is particularly relevant because it may address several of the specific barriers identified in this study. Compact low-field and ultra-low-field systems represent one important approach: they can reduce requirements for power, cooling, shielding, and dedicated installation space, and several designs have been developed specifically to improve portability and reduce infrastructure demands (Liu et al., 2021; Geethanath and Vaughan, 2019; Cooley et al., 2021; Arnold et al., 2023; Obungoloch et al., 2023; Sarracanie et al., 2015; Lother et al., 2016; Cooley et al., 2015; Zhao, Ding, et al., 2024; O’Reilly and Webb, 2022; Chang et al., 2006; Halse et al., 2006; Sheth et al., 2022; Vaughan et al., 2016). These characteristics could make deployment outside conventional tertiary radiology settings more feasible, including in locations where electrical and physical infrastructure are limited.

However, lower infrastructure requirements do not resolve many of the barriers that determine whether an imaging service is sustainable. Accessible MRI systems still require trained personnel, image interpretation, quality assurance, maintenance, and access to technical support and replacement components (Geethanath and Vaughan, 2019; Arnold et al., 2023; Obungoloch et al., 2023; Deoni et al., 2022). In a setting with limited local service capacity, these operational requirements may ultimately be as important as scanner purchase price. Accessible MRI should therefore be viewed as a means of reducing selected infrastructure barriers, not as a technological solution to the broader constraints affecting neuroimaging access.

Its clinical role also needs to be defined according to local diagnostic needs. Portable and low-field MRI has shown potential for evaluating ischemic stroke and other acute neurological conditions, including bedside assessment of critically ill patients and intracranial mass effect (Parasuram et al., 2023; Yuen et al., 2022; Beekman et al., 2022; Sheth et al., 2020; Sheth et al., 2022). Studies have also demonstrated the feasibility of detecting intracerebral hemorrhage at low field, although hemorrhage detection remains an important limitation and CT continues to offer important advantages for rapid acute evaluation (Mazurek et al., 2021; Mazurek et al., 2023). Advances in image reconstruction, super-resolution, and electromagnetic interference suppression may further improve low-field performance (Lau et al., 2023; Koonjoo et al., 2021; Srinivas et al., 2022; Zhao, Xiao, Hu, et al., 2024; Zhao, Xiao, Liu, et al., 2024), but diagnostic performance remains application-dependent.

These considerations suggest that the most appropriate role for accessible MRI in Chad would likely be complementary to existing CT services rather than a replacement for them. The relevant benchmark is not whether low-field MRI can reproduce all capabilities of a 1.5 T or 3 T system, but whether it can answer clinically important questions that currently go unanswered at a cost and level of complexity that can be sustained locally. This shifts the focus from field strength alone to context-specific clinical value and provides a more practical framework for evaluating whether accessible MRI could meaningfully expand neuroimaging access in Chad.

### 4.3 Implementation considerations and priorities

The potential benefits of accessible MRI will depend heavily on how the technology is implemented. Rather than deploying scanners broadly at the outset, a staged approach emphasizing local acceptance that begins at selected sites with substantial neurological demand and sufficient clinical support would allow feasibility, diagnostic value, and operational requirements to be assessed before wider expansion. Experience with low-field MRI development in Africa and broader frameworks for sustainable MRI access similarly emphasize that equipment selection should be integrated with local infrastructure, workforce, financing, and long-term technical support (Obungoloch et al., 2023; Anazodo et al., 2023).

Workforce and technical capacity are likely to remain major constraints even if scanner infrastructure is simplified. Expansion of imaging services will require not only trained operators and interpreting physicians, but also personnel capable of quality assurance, routine maintenance, and first-line troubleshooting. Training approaches tailored to underserved and rural settings may help address this gap (Kawooya, 2012), while teleradiology could extend access to specialist interpretation where on-site expertise is limited (Tahir et al., 2022; Essop and Kekana, 2020). However, these approaches themselves depend on reliable digital infrastructure, appropriate workflows, and local personnel capable of integrating remote expertise into clinical care.

Long-term sustainability should therefore be evaluated alongside image quality and acquisition cost. Systems that reduce power requirements, simplify maintenance, and permit greater local technical involvement may offer advantages even if they are not the least expensive to purchase (Obungoloch et al., 2023; Rezaei et al., 2022). Improvements in distributed or renewable energy infrastructure may also help support imaging services in areas with unreliable electricity (Byaro and Mmbaga, 2022). Initial implementation studies should assess not only diagnostic performance, but also scanner uptime, maintenance requirements, operating costs, workforce needs, patient accessibility, and effects on clinical decision-making. For deployment outside conventional radiology environments, ethical and logistical considerations associated with community-based imaging should also be addressed (Shen et al., 2024).

Overall, implementation should be guided by sustainable service delivery rather than equipment deployment alone. Accessible MRI may lower some barriers to establishing imaging services, but its long-term impact will depend on whether the technology can be integrated with local clinical pathways, personnel, maintenance capacity, and referral networks, as well as existing healthcare financing, regulatory, reimbursement, and public health policy frameworks.

### 4.4 Limitations and future directions

Several limitations should be considered when interpreting these findings. First, hospital data were available from a limited number of institutions in N’Djamena and were not uniform across hospitals, diagnoses, or reporting periods. Differences in record-keeping and data-sharing practices prevented construction of a comprehensive national dataset. The reported hospitalization counts should therefore be interpreted as selected institutional data rather than estimates of population-level incidence or prevalence. Because specialized neurological services are concentrated in the capital, the available data may also inadequately represent disease burden and barriers to care in rural and remote regions.

Second, contextual information from neurologists, radiologists, and hospital personnel was obtained through open professional communications rather than structured interviews or surveys. These observations provided important practical context but should not be interpreted as a systematic assessment of provider experiences or opinions. Future studies incorporating structured input from healthcare professionals and patients could more comprehensively characterize barriers to neuroimaging access.

Third, comparisons of MRI availability across African countries are limited by heterogeneity in the year, source, and scope of the available data. For some countries, reported scanner counts represent only public- or private-sector equipment. **Table 2** therefore provides a broad comparison of MRI availability rather than a precise contemporaneous ranking of national imaging capacity. More standardized reporting of scanner numbers, operational status, geographic distribution, and imaging workforce would improve assessment of neuroimaging capacity across the region.

Finally, accessible MRI has not yet been prospectively evaluated within the Chadian healthcare system. Although previous work demonstrates the feasibility of developing and deploying low-field MRI in resource-constrained settings (Obungoloch et al., 2023; Anazodo et al., 2023), the optimal technology, clinical applications, and deployment model for Chad remain uncertain. Future pilot studies should therefore evaluate not only diagnostic performance, but also reliability, maintenance requirements, workforce needs, operating costs, patient accessibility, and integration with existing clinical pathways. Community-based deployment would additionally require attention to the ethical and logistical considerations associated with imaging outside conventional radiology environments (Shen et al., 2024).

## Data Availability

All data produced in the present work are contained in the manuscript.

## Funding Declaration

The U.S. component of this work was supported in part by U.S. National Institutes of Health grants R01HD093578, R01HD099846, and R01HD110152 (PI: A.V.D.K.).

## Acknowledgements

We are grateful to the following individuals from around the world for their insightful discussions and expertise: Drs. Bruce Rosen, David Rosman, Rory Cochran, Jad Husseini, Susie Huang, Ernesta Meintjes, Ethar Khalil Ibrahim, Kamis Dakisia, Desire Nalire, Nderbe Melom, Ousman Alkher, Madjirabe Christian, Kebede Beshah, Udunna C. Anazodo, Abdelmadjid Abderahman, Saleh Hassan, and Ms. Debbie Oubre. We also thank Deoutol Alladjaba and Fatchou Soukassia Appolinaire for their assistance in collecting data from major hospitals in Chad.

## Author Contributions

I.H., J.Y., J.L.A., and A.V.D.K. conceptualized the study. I.H. and J.Y. designed the study and drafted the manuscript. F.S. established contact with the Chadian neurologists and contributed to data collection from hospitals in Chad. M.Z. conducted the technical analysis of MRI systems. E.B. developed the policy recommendations and evaluated the study’s limitations. All authors were involved in information collection and consultation, participated in manuscript revision, and approved the final version.

## Competing Interests

The authors declare no competing interests.

## Data Availability Statement

All data generated or analyzed during the study are included in this published article. No identifiable patient-level data were accessed or used, and no data were collected through formal surveys or structured interviews.

## Notes

### Competing Interest Statement

The authors have declared no competing interest.

### Author Declarations

All data generated or analyzed during the study are included in this published article. Data sources include publicly available reports, government and NGO websites, published literature, and open professional communications with Chadian neurologists and radiologists. No data were collected from human subjects or through formal surveys.

